# Evaluating a novel, integrative dashboard for health professionals’ performance in managing deteriorating patients: *quality improvement project*

**DOI:** 10.1101/2022.06.28.22276946

**Authors:** Baneen Alhmoud, Daniel Melley, Nadeem khan, Timothy Bonnici, Riyaz Patel, Amitava Banerjee

## Abstract

**Background:** The quality of recording and documentation of deteriorating patient management by health professionals has been challenged at health system level during the COVID-19 pandemic. Non-adherence to monitoring and escalation guidelines and poor documentation increases risk of serious adverse events. Electronic health record (EHR)-integrated dashboards are real-time auditing tools of patients’ status and clinicians’ performance, but neither the views nor the performance of health professionals have been assessed, relating to management of deteriorating patients..

**Objective:** To develop and evaluate a real-time dashboard of deteriorating patients’ assessment, referral, and therapy by examining the perception of the dashboard and the performance of nurses and physicians.

**Settings:** Five academic hospitals in the largest NHS trust in the UK (Barts Health NHS Trust).

**Intervention:** The dashboard was developed from EHR data to investigate patients with NEWS2>5, assessment, and escalation of deteriorating patients. We adopted the Plan, Do, Study, Act model and followed the SQUIRE framework to evaluate the dashboard.

**Design:** Mixed methods: (i) Virtual, face-to-face, key informant interviews and (ii) Retrospective descriptive EHR data analysis to measure performance change over time.

**Results:** We interviewed 3 nurses (2 quality and safety and 1 informatics specialists). Key themes were: (1) participants perceived the dashboard as a facilitator for auditing NEWS2 recording and escalation of care to improve clinicians practice; (2) There is a need for guiding clinicians and adjusting data sources and metrics which could enhance the functionality and usability. From EHR (2019 to 2022) data analysis showed: (1) NEWS2 recording has gradually improved in the implementation and evaluation phases (May 2021 to Apr 2022) from 64% to 83%; (2) Referral and nurses’ assessment forms completion increased (n: 170 to 6800 & 23 to 540, respectively).

**Conclusion:** The deterioration dashboard is an effective real time data-driven method for improving the quality of managing deteriorating patients. Improving the dashboard by integrating multiple health systems, a wider analysis of further NEWS2 and escalation of care metrics, clinicians’ learning of digital solutions will enhance functionality and experience, potentially boosting its value. There is a need to examine the generalizability of the dashboard through further validation and quality improvement studies.

## Introduction

The Covid 19 pandemic has taken its toll on health care services globally. The escalating pressure has significantly raised the surge in deteriorating patients and the need to escalate their care(1). There was an increase in daily tasks for nurses, physicians and rapid response teams to cope with the COVID-19 strain(1). Clinicians’ practice can be adversely affected by the increased patient to staff ratios (2), the complexity of patients’ clinical care(3), and the ongoing pandemic impact on the healthcare service and individual staff (4). The quality of clinicians’ assessment, documentation, and timely referral for escalation can suffer.

Early warning scores (EWS) are widely implemented predictive tools to detect deterioration in an early stage of critical illness. Their performance has been variable (5), and their effectiveness is subject to multielement in the clinical settings(6,7). EWS performance in detecting critical events is not only related to the score’s sensitivity. Nurses’ adherence to recommended monitoring and escalation guidelines and physicians’ compliance with critical events and sepsis assessment may correlate with the outcomes studied in EWS validation (8). Common problems found in clinicians’ behaviour toward EWS protocol include non-compliance with recommended monitoring frequency, notification of doctors when indicated by EWS, or timely response of doctors and CCRT (8,9). Therefore, serious adverse events (SAE) occur due to misclassification of patients and poor allocation to critical care despite the implementation of EWS and escalation guidelines. Along with established implementation and validated performance of EWS, human factors are vital for the success of EWS application.

Real-time auditing can be an effective method to detect the roots of clinicians’ adherence defects. With the availability of Electronic health records (EHR) systems, a representative, generalisable dataset can be captured and analysed at scale via integrated EHR dashboards in a constructed, organised form (10). Healthcare dashboard is an electronic analytics tool to monitor healthcare key performance indicators(KPI) by displaying outcomes, auditing progress, identifying deficiencies, and manage professional and clinical activities in healthcare organisations (11–13). Digital dashboards systems capture EHR data to generate information on the healthcare system, individual professionals’ performance, and the patient’s prognostic status. Prompt, concise and context-specific display of the performance provides analysis of hospital and patients status, facilitating clinical decision making and quality improvement (14,15). For example, the NHS Pathways of Coronavirus Triages and Activity dashboards in NHS hospitals are examples of live data visualisation for the public information at an organisational level (16). Although dashboards have proven efficiency in providing real-time information for hospital management and stakeholders (14,17,18), functionality is limited when addressing performance issues from patient chart data. Logging into each patient’s chart several times during the day for specific information is time-consuming and problematic, i.e. completion of EWS recording in a day shift. Healthcare dashboards are concise, time-saving, and intuitive tools for up-to-date assessment and escalation auditing of deteriorating patients.

### Problem

Despite the widespread implementation and expenditure of EHRs in healthcare settings, secondary use of data for improving quality and safety is limited. The full potential benefits from EHRs data are far from realisation currently despite massive efforts of investment in health technology(19). Challenges such as the conflicts between public interest, individual patient safety, and optimising application of health information systems, have restricted the potentials of data use (20,21). With the growing integration of EWS into EHR, a significant amount of data for critically ill patients are available (22), but not yet utilised.

Furthermore, COVID-19 has affected the quality of clinicians’ routine practice and hindered tasks that could be regularly and efficiently carried out prior to the pandemic (23). As a result, appropriate, timely management of acutely ill patients declined(24,25). Escalating care of an acutely ill patient must follow timely and careful assessment, then communicating the evaluation to the designated critical care professional for further intervention. Errors in detecting worsening of the condition and failure to communicate or intervene can hamper escalation of care and negatively impact clinical outcomes and NEWS2 performance. Noncompliance with escalation protocols or recommended documentation guidelines may result in serious healthcare errors (26). The Record Management Code Practice 2021 provides a framework to guide organisational and individual responsibilities when managing patient records (26). Auditing is an integral part of healthcare records policy and guidelines to assess the standard achieved in records and find areas needing improvement for health data and staff (27).

In Barts Health NHS Trust, the National Early Warning Score (NEWS2) has been nationally endorsed and implemented (28). A digital transformation took place in Barts hospitals by shifting NEWS2 recording into digital format and automating routine monitoring, hoping to increase accuracy of information if the escalation protocol is optimally followed. However, implementing digital NEWS2 requires complete and updated her, which has been more difficult in the COVID context at Barts and other hospitals. Local audits in the trust showed non-adherence to routine monitoring and recording, e.g. in 2019-2020 NEWS2 status was incomplete and not in line with guidelines in approximately a quarter of the patients’ population (Complete vitals: 61-79%, 75-83%, 84-87%, 83-91% & 79-86% from September 2019 - January 2020 in Newham, Royal London, St Bartholomew’s, and Whipps Cross hospitals, respectively) (Appendix 1).

A dashboard integrated in EHR would allow performance of health professionals, data quality patient care to be monitored and improved by facilitating timely health resources management and support informed clinical decision making. We therefore conducted a quality improvement study to evaluate deteriorating patient dashboard and provide evidence for an exemplary quality of care for other healthcare settings.

### Aims

To create and evaluate an EHR data-driven dashboard of real-time assessment of deteriorating patients and escalation of care. We aim to evaluate the dashboard through PDSA cycles, including:

1. Examine the views on the dashboard and areas that need improvement from key users’ perception; and implement necessary actions for development.
2. Evaluate the performance of nurses and physicians in the stages of managing deteriorating patients on all trust levels through historical tracking of data.

### Ethical consideration

The study is registered and approved by the Health Research Authority (HRA) and Health and Care Research Wales (HCRW) and sponsored by University College London (UCL). The Stanmore Research Ethics Committee (REC) in London approved conducting the study; reference number: 20/PR/0286.

## Methods

### Context

The dashboard was implemented in the largest NHS trust in the UK (Barts Health NHS Trust) in five academic hospitals: Mile End Hospital, Newham University Hospital, Royal London Hospital, Whipps Cross University Hospital and St Bartholomew’s Hospital. The dashboard was developed by creating Vitals data table, then transformed into a thorough and more robust data visualisation of NEWS2, assessment and escalation of deteriorating patients via Qlik Sins. Development was led by DM and executed by NK. Data of around 1.2 million recordings of 110,000 admissions from August to October 2020 were extracted from EHR (Cerner Millennium®) (Figure 1 & appendix 2).

**Figure 1.**
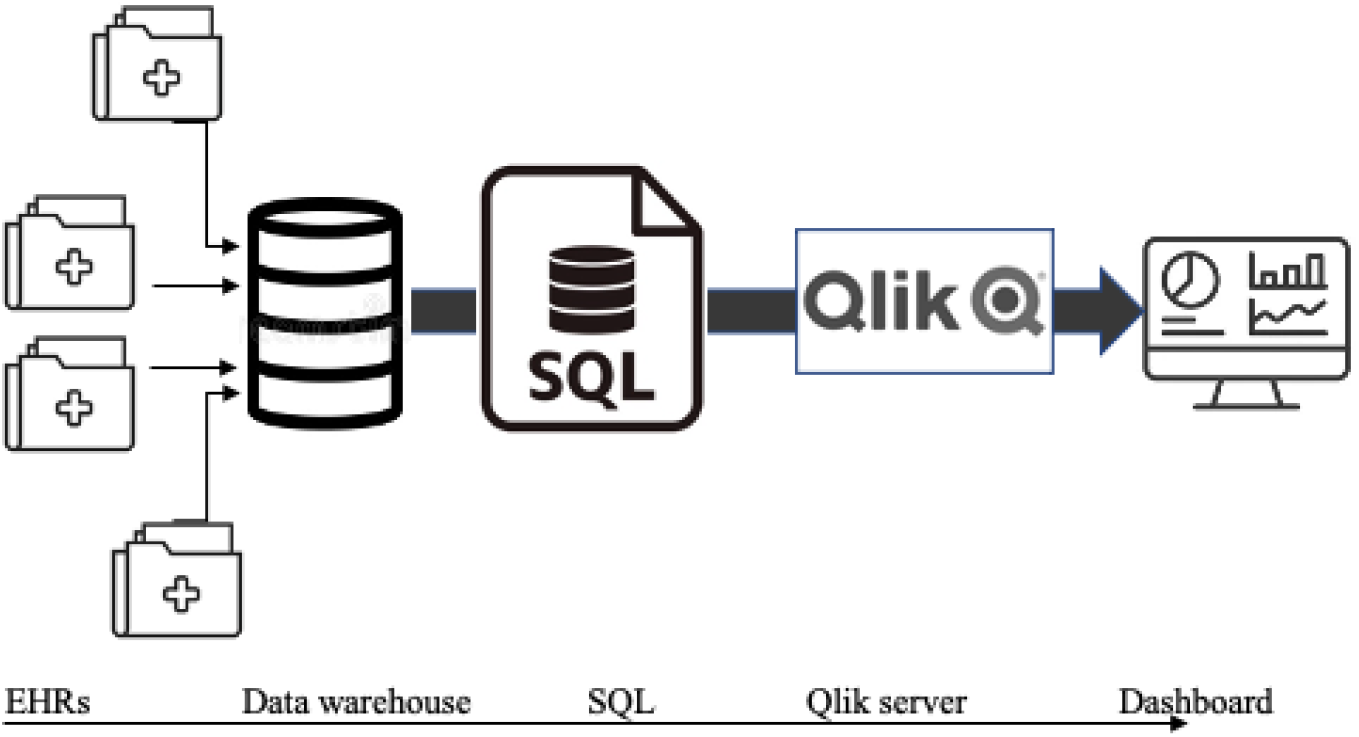
Illustration of deteriorating patient’s dashboard development from patients EHRs data.

The user interface includes live and accumulative data of patients with high NEWS2, and performance tracking of stages in deteriorating patients’ management by nurses and physicians on all trust levels. Health professionals’ performance is measured by the completion of the assessment, escalation of care, and sepsis treatment (Figure 2)

**Figure 2.**
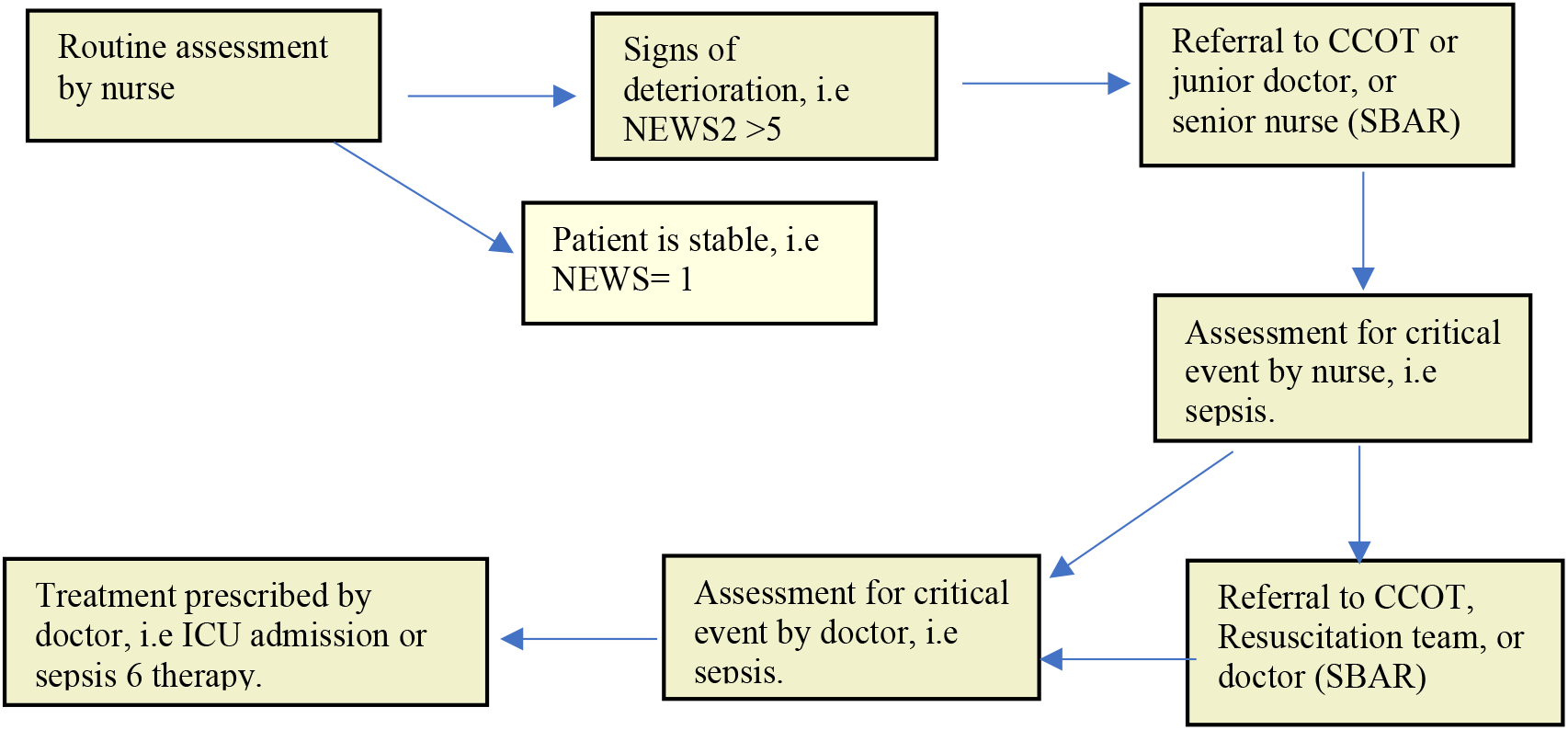
Escalation of care assessment flowchart. Abbreviations: CCOT: critical care response team, SBAR: Situation-Background-Assessment-Recommendation tool for communication between health care team.

### Intervention

We adopted the PDSA (Plan-Do-Study-Act) (29) model to examine the objectives of creating the dashboard and evaluating clinicians’ performance in the deteriorating patient management cycle. The PDSA model involves:

- Plan: plan the test, intervention, or observation, including a method for collecting data.
- Do: conduct the intervention on a small to a bigger scale.
- Study: analyse the data and study the results.
- Act: refine the change based on what was learnt from the best.

### PDSA cycle 1

We planned to implement the dashboard and evaluate the performance from the perspective of key people in management who utilised it for auditing. The dashboard was initially launched in May 2021, and several improvements to the metrics and filters to serve the function until the version which we investigated in September 2021. The dashboard was introduced to ward managers, quality improvement, and patient safety teams. The improvement plan was mapped out for the involved teams by informing them of the release of the dashboard and its objectives and functions. The guidelines for the utilisation of the dashboard were not developed fully yet; therefore, they were not disseminated to ward managers. By the end of this phase (October 2021), we interviewed three staff who took part in the initial roll-out: senior nurse for quality and safety, nursing informatics specialist, and patients safety practitioner. The information collected allowed us to create the next PDSA cycle to improve the effectiveness of the dashboard for a wider cohort of users.

### PDSA cycle 2

In this phase, the Electronic Prescribing for Medicine Administration (EPMA) implementation has already taken place. A how-to guide was developed to educate users on effectively making the most of the dashboard. Due to the EPMA’s role in handling drug prescribing data, data for this metric were missing from EHRs and have to be extracted from EPMA. After refining the dashboards and releasing the guidance, managers were encouraged to view and report information from the dashboard, and nurses and doctors were informed of areas in practice that needed adjustments. Data from the dashboard user interface were assessed to evaluate changes through time in deteriorating patients’ management. There was a plan to integrate EPMA data into the dashboard in the coming stage and provide further valuable data, i.e. time of treatment prescribing.

### Measurements

We conducted individual interviews to evaluate the perception on the dashboard. Interview questions were created to gather qualitative data and adapted from a previous evaluation of dashboards of ward specific performance (12) and aligned with the Technology Acceptance Model framework (TAM)(13) (Figure 3). The key questions were on the perceived benefit, usability, the intention to use and the actual functionality, and desired adjustments needed to improve the dashboard (Appendix 2). A purposive sample of key users, as guided by DM, was interviewed by BA.

**Figure 3.**
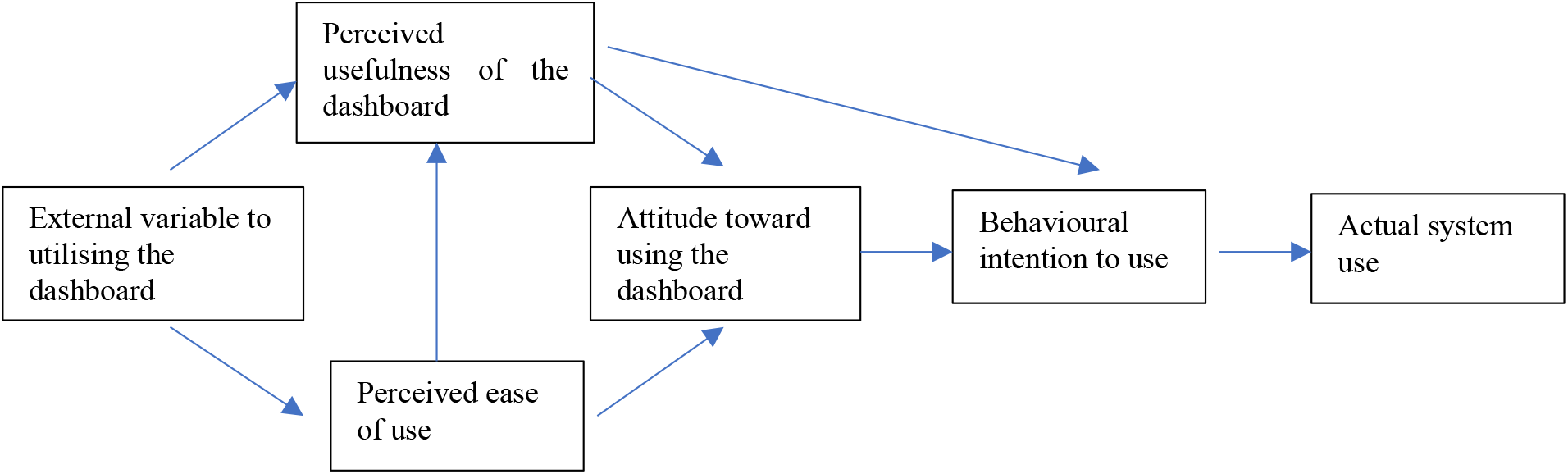
Illustration of the Technology Acceptance Model (TAM) to examine the perception of the deterioration dashboard

A before-and-after data evaluation was run to assess the change in performance in the measured metrics. The period was divided into five phases to interpret the improvement in recordings and forms completion. Phases were as follows: pre-EHR integration (August-November 2019), post-EHR integration (December 2019-September 2020), automation period (October 2020-April 2021), implementation period (May -September 2021), and post-feedback (October 2021-April 2022). Due to the potential effect of skewing doctors’ assessment and treatment data caused by shifting entry from EHRs to EPMA, we compared the two metrics between the Dashboard period (May to August 2021) with post-EPMA rollout (September 2021 to April 2022).

### Analysis

Interviews were audio-taped and transcribed, and qualitative data were analysed using NVivo software following a content analysis approach by B.A. The method is suitable for analysing interviews through systemic coding of transcriptions to indicate the presence of meaningful content related to the evaluated domains. Data analysis was iterative, where content was coded, grouped to form subcategories, and then into themes that represent the topics creating the focus of the evaluation. The transcripts and analysis were checked independently as developed by two researchers to ensure rigour.

Descriptive analysis was done on the data collected from the dashboard using the R programme. Pearson’s chi-square test was used to compare periods identified for NEWS2 recording and forms completion. A *p-*value of <0.05 is considered statistically significant.

## Results

Three participants were interviewed. Participants expressed their perceived advantages and usability of the dashboard for escalation of care, auditing NEWS2 recording and forms completion, and areas in need of improvement. The interviews data content formed two elements: (i)dashboard function, and (ii) obstacles and improvement. From the descriptive data, we found a gradual improvement in NEWS2 and forms compliance by nurses and doctors

### Dashboard function

The dashboard is perceived was a tool for quality improvement. There was an agreed perception of its analytics function on individual nurses’ and doctors’ performance in the escalation of the care process. Primary auditors are ward managers, senior nurses, and quality improvement officers. They could view periods and specific ward improvements and where the trend is declining; in patients’ status and clinician’s practice; to analyse the reasons for changes to planning for enhancement.

The dashboard helped nurses and doctors chase the escalation of deteriorating patients from the auditing function by monitoring NEWS2, SBAR referral, and assessment completion to push for a better result.

Another benefit found was the attention to one’s performance due to being tracked in real time. It was reported that staff were becoming more interested in completing the forms and monitoring within the time frame and how properly the documentation in return to the observed numbers of deteriorating patients scoring high NEWS2.

### Obstacles and improvements

Participants reported issues in lack of engagement by managers in relation to difficulty in usability, such as locating and navigating it properly. It was suggested to improve in the dashboard function and utility. They recommended comprehensive showcasing of the dashboard for clinicians to understand the benefits and purpose. Participants expressed the need for a clear guide for utilisation to encourage clinicians make the most of the dashboard confidently. In addition, it was suggested for databases to be stored in a unified standard system to facilitate data extraction and query writing and explore the possibility of creating more functions from the health systems. Several additions were recommended by participants to enhance the role of the deterioration dashboard, including additional assessment metrics, sepsis diagnosis and treatment time, and monitoring wrist bands scanning for ID confirmation.

### Compliance measure

The audit showed poor compliance with vital signs and NEWS2 recording in the baseline period (64%), then improved gradually after the EHR integration of NEWS2 (81.5%), followed by an increase after automating vitals monitoring and dashboard implementation (85 & 83%, respectively). Patients with high NEWS2 reached a peak between April and May 2020 and in January 2021 (∼25%) when the first and second waves of COVID-19 occurred. Complete referral and nurse assessment forms were boosted after dashboard implementation (n: 170 to 6800 & 23 to 540, respectively). The screening and prescribing by doctors improved in the first dashboard phase (n: 22 to 36 &15 to 26, respectively), then had a sudden drop (n: 8 &6, respectively) after EPMA became the data entry point (Table 1 and Figure 4).

**Table 1.** Definitions of terminologies.

| Definitions |  |
| --- | --- |
| Dashboard | Electronic analytics tool to monitor key performance indicators (KPI) by displaying outcomes, progress, deficiencies in an organisation(13,16) |
| Automated monitoring | Integration of patients routine monitoring and EHRs by transmitting measurements directly from monitoring machine to patients' charts and continuously calculating and updating NEWS2 (30,31). |
| Electronic Prescribing for Medicine Administration (EPMA) | Electronic system to facilitate the communication of a prescription, aiding the choice, administration, and supply of a medicine through decision support. (32) |
| Situation, Background, Assessment and Recommendation tool (SBAR) | A communication tool including, Situation, Background, Assessment, Recommendation; for structuring conversations between doctors and nurses about situations requiring attention (33) |
| Sepsis 6 | A structured care bundle for patients with sepsis including blood cultures, check full blood count and lactate, IV fluid challenge, IV antibiotics, monitor urine output and give oxygen (34). |

**Table 1:** Dashboard metrics trend measured in different phases.

| Measure | Routine monitoring phase. (Aug-Nov 2019) (min-max, median) | EHRs-EWS phase. (Dec 2019-Sep 2020). (min-max, median) | Automation phase. (Oct 2020-Apr 2021). (min-max, median) | Implementation phase (May-Sept 2021). (min-max, median) | Evaluation phase (Oct 2021-Apr 2022). (min-max, median) | P value |
| --- | --- | --- | --- | --- | --- | --- |
| Vitals (%) | 59-72, 64 | 74-84, 81.5 | 84-86, 85 | 81-84, 83 | 81-83, 82 | 0.01538 |
| NEWS>5 (%) | 5-6.5, 6 | 5-17.5, 6 | 4-20, 11.5 | 5.5-7, 6 | 5-7, 6.5 | 0.4978 |
| SBAR (n) | - | - | 100-230, 170 | 220-3100, 650 | 3800-8300, 6800 | 0.3792 |
| Nurse screening (n) | - | - | 13-94, 23 | 36-250, 75 | 250-750, 540 | 0.3792 |
|  |  |  | Automation phase. (Oct 2020-Apr 2021). (min-max, median) | Implementation phase (May-Aug 2021) (min-max, median) | EPMA phase (Sept 2021-Mar 2022) (min-max, median) | P value |
| Doctor screening (n) | - | - | 10-82, 22 | 25-60, 36 | 3-14, 8 | 0.03528 |
| Sepsis 6 (n) | - | - | 8-64, 15 | 19-47, 26 | 1-12, 5.5 | 0.2415 |

Note: Each number is presented as minimum to maximum and median.
Abbreviations: Vitals: vital signs recordings; NEWS2: Updated National Early Warning Score; SBAR: situation, background, assessment, recommendation; Sepsis 6: clinical care bundle for sepsis management in the first hour of diagnosis.

**Figure 4.**
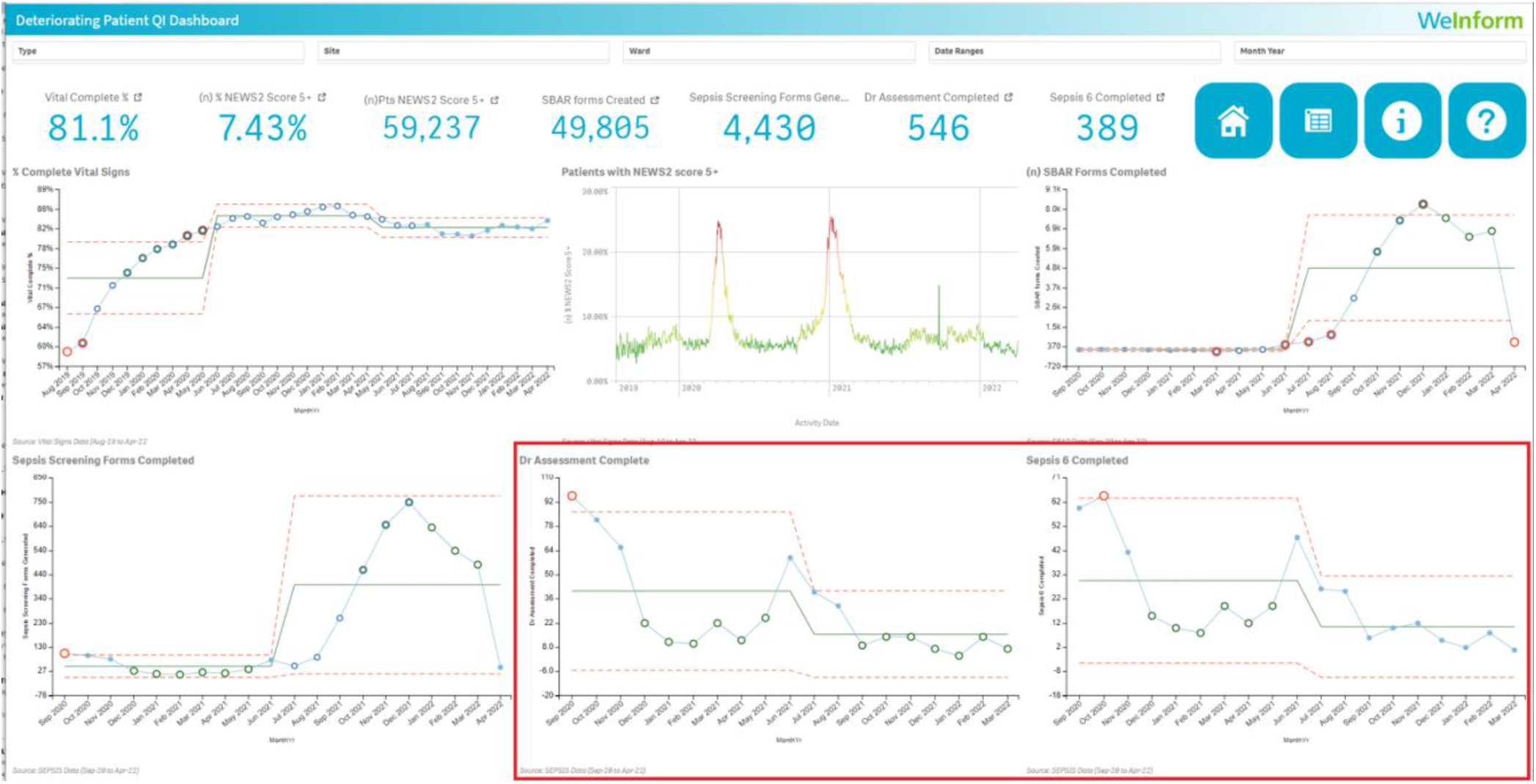
Snapshot of front page of deteriorating patients’ dashboard from August 2019 to March 2022. Abbreviations: SBAR: esclation of care handover tool (situation, background, assessment, and recommendation); Sepsis screening forms: assessment of sepsis forms by nurses; Dr Assessment complete: sepsis screening by doctors; Sepsis 6: prescribing sepsis 6 bundle therapy. Colour codes: orange: upper and lower control limits and outliners; light blue: complete; dark blue: trends; green: central lines; graphs with red frame represent metrics that will be adjusted in the next stage.

## Discussion

We report the experience of development and implementation of an EHR-integrated dashboard for NEWS2 and escalation of care auditing in cardiac specialist teaching hospitals. This novel and unique initiative focused on monitoring and improving early recognition and expediting shared care by nurses and physicians. Our data visualisation is a practical and effective way for performance tracking; and, therefore, promoting timely NEWS2 recording and forms completion for improved escalation of care. We had three main findings. First, participants perceived the dashboard as an excellent auditing tool that improves deteriorating patients’ care. Second, some improvements are needed to enhance its functionality and user experience. Third, historical data have shown an increase in clinicians’ compliance with documentation and escalation protocol that may have responded to monitoring individual and unit performance and, therefore, driving quality improvement.

We found our results consistent with previous findings confirming the effectiveness of dashboard analytics in evaluating clinical performance (35,36). It has been shown that dashboards are visual tools to examine the programme or protocol’s effectiveness in meeting the objectives (16). The perceived individual staff monitoring and improving own actions came supporting to previously highlighted impact of dashboards on nurses’ awareness. Nurses’ attention to the ward’s accomplishments affected patients’ outcomes and gave them a sense of control and satisfaction with their achievements (13,17,36). Investigating the deteriorating patient management provides on-time tracking of escalation of care steps and NEWS2 recording to have a transparent understanding of its predictive performance in the care settings. Display and use of performance data are keys to identifying areas of strengths and weakness for quality improvement.

For a dashboard system to be widely used, it must be an easy, user-friendly, and intuitive system. Our results indicate the need for clinicians to learn about digital tools’ usability, as the previous implementation of digital health systems showed the demand for embedding technology training and education (37,38). In addition, expanding the structure and functionality through integrating different data sources and refining the design is a substantial gain. Previous health dashboards where EHRs are combined with other systems, such as PACS, have given professionals a broader view and knowledge of patients’ health status (14). By integrating multiple digital systems, there is a potential for great use of information to understand, research, and explain unclear data of one system by the other. The reduction in doctors’ assessment and Sepsis 6 was due to the data entry shift from EHRs to EPMA. In the trust, EPMA is believed to add the benefit of providing more true, timely treatment information for auditing. EPMA integration can provide data on prescribing antibiotics as per the recommended protocol, therefore, will be conducted in the next phase. Other data resources could also enhance and maximise the functionality, such as COVID-19 patients’ data, resources tracking such as ICU beds and Critical Care Outreach (CCOT) staff, and timeliness of escalation of care steps data. Data sources addition could help doctors, nurses and managers organise the treatment plan promptly for clinicians’ and patients’ benefit.

From time series analysis, we interpreted a positive change in NEWS2 recording and formed completion post dashboards intervention and potential for a further improvement as quality is monitored. In the current integration of EWS into EHRS, displaying the real-time score and generating alerts of EWS, like Modified Early Warning Score (MEWS), Paediatric EWS (PEWS), and NEWS, has shown several advantages in different care settings (39–41). It allowed for a real-time prediction of critical events associated with reduced hospitalisation costs and, more importantly, is believed to be a keystone for safe practice. In addition, dashboards have been increase implemented in healthcare and supported healthcare services, such as communicating patient-reported and clinical data in cystic fibrosis (42). Ward patient status and clinician performance auditing represent a modern method of quality improvement in the digital clinical environments to be promoted and examined widely.

Several additional characteristics could enhance the function of dashboard auditing deterioration management. Producing a live NEWS2 score for patients would add an advantage to estimating validation if analysed on wards and specialities level. In addition, alerting the first-line responders to escalation, including the CCOT team and bedside nurses, would benefit managers in tracking the escalation and whether it occurred due to the score, clinicians’ observation or the two combined. Expanding the functionality guided by the escalation of care and early warning scores protocols would be needed to enhance the performance impact. However, further studies need to evaluate the effect of advancing the dashboard in the coming stages from a user perspective and the extent to which it can positively impact clinical care outcomes and work performance.

### Limitations

The dashboard was developed and tested at the time of the COVID-19 pandemic. This factor could have affected the engagement of key users and, therefore, the response to utilising the dashboard for auditing. Another disadvantage is the small team of clinical informatics that developed the dashboard during the pandemic pressure, which might have resulted in delays for further adjustments and applying a third PDSA cycle. We interviewed three health professionals during the examined phases; feedback from ward managers, nurses, and doctors will show a better view of its validity for monitoring escalation of care. Furthermore, we did not verify the generalizability of the dashboard system in other trusted hospitals. Studies need to examine its feasibility and usefulness in different hospitals with differing structures and patients’ population.

## Conclusion

On-time data visualisation of deteriorating patient care is an effective and efficient method for establishing quality improvement. The deteriorating patient dashboard facilitated timely investigation and improvement of the practice of assessment and treatment from key users’ perspectives and performance analysis over time. Evaluating adherence to NEWS2 recording and escalation of care protocol can help clarify EWS validation where it is implemented.

Advancing health dashboards by facilitating multiple health systems integration and clinicians learning digital health solutions will enhance dashboard functionality and improve user experience. In addition, functionality could be upgraded by analysing further NEWS2 and escalation of care protocol metrics and times; promoting live and historical data value. There is a need for further validation and quality improvement studies to verify the generalizability of the dashboard system in different settings.

## Supporting information

Squire framework

## Data Availability

All data relevant to the study are included in the article or uploaded as supplemental information

## Other information

### Ethics statement

The study is approved by HRA and HCRA, REC reference: 20/PR/0286

### Patient consent for publication

Not applicable.

### Participants’ consent

Informed consent is obtained prior to interviews.

### Contributors

DM has developed the project plan, and NK executed the development with guidance from DM. BA evaluated the dashboard, including interviews and analysis, with the guidance of AB. BA wrote the manuscript, and all authors contributed to the finding’s interpretation and revision of the manuscript.

### Data sharing statement

All data relevant to the study are included in the article or uploaded as supplemental information.

### Twitter

@BaneenAlhmoud, @DanBartsICU, @amibanerjee1, @TimBonnici, @DrRiyazPatel.

