## Supplementary material for "Evaluating a novel, integrative dashboard for health professionals’ performance in managing deteriorating patients: *quality improvement project*": Squire framework

| **Research and reporting methodology** |  | |
| --- | --- | --- |
| Revised **Standards for QUality Improvement Reporting Excellence** (**SQUIRE 2.0**) publication guidelines |  | |
| **Notes to authors** |  | |
| ▸ The SQUIRE guidelines provide a framework for reporting new knowledge about how to improve healthcare. |  | |
| ▸ The SQUIRE guidelines are intended for reports that describe system level work to improve the quality, safety and value of healthcare, and used methods to establish that observed outcomes were due to the intervention(s). |  | |
| ▸ A range of approaches exists for improving healthcare. SQUIRE may be adapted for reporting any of these. |  | |
| ▸ Authors should consider every SQUIRE item, but it may be inappropriate or unnecessary to include every SQUIRE element in a particular manuscript. |  | |
| ▸ The SQUIRE glossary contains definitions of many of the key words in SQUIRE. |  | |
| ▸ The explanation and elaboration document provides specific examples of well-written SQUIRE items and an in-depth explanation of each item. |  | |
| ▸ Please cite SQUIRE when it is used to write a manuscript. |  | |
| **Text section and item name** | | **Page/line no(s).** |
|  | | **info is located** |
| **Title and abstract** | |  |
| 1. **Title** | | Page 1 |
| Indicate that the manuscript concerns an initiative to improve healthcare (broadly defined to include the quality, safety, effectiveness, patient-centredness, timeliness, cost, efficiency and equity of healthcare). | |  |
| 2. **Abstract** | | Page 2 |
| a. Provide adequate information to aid in searching and indexing. | |  |
| b. Summarise all key information from various sections of the text using the abstract format of the intended publication or a structured summary such as: background, local problem, methods, interventions, results, conclusions. | |  |
| **Introduction: Why did you start?** | |  |
| 3. **Problem description** - Nature and significance of the local problem. | | Page 3 and4 |
| 4. **Available knowledge** - Summary of what is currently known about the problem, including relevant previous studies. | | Page 3 |
| 5. **Rationale** - Informal or formal frameworks, models, concepts and/or theories used to explain the problem, any reasons or assumptions that were used to develop the intervention(s) and reasons why the intervention(s) was expected to work | | Page 3 and 4 |
| 6. **Specific aims** - Purpose of the project and of this report. | | Page 4 |
| **Methods: What did you do?** | |  |
| 7. **Context** - Contextual elements considered important at the outset of introducing the intervention(s). | | Page 5 and  Figure 1 |

| 8. **Intervention(s)** | Page 5 |
| --- | --- |
| a. Description of the intervention(s) in sufficient detail that others could reproduce it. |  |
| b. Specifics of the team involved in the work. |  |
| 9. **Study of the intervention(s)** | Page 5 and 6 |
| a. Approach chosen for assessing the impact of the intervention(s). |  |
| b. Approach used to establish whether the observed outcomes were due to the intervention(s). |  |
| 10. **Measures** | Page 6 and Figure 2 |
| a. Measures chosen for studying processes and outcomes of the intervention(s), including rationale for choosing them, their operational definitions and their validity and reliability. |  |
| b. Description of the approach to the ongoing assessment of contextual elements that contributed to the success, failure, efficiency and cost. |  |
| c. Methods employed for assessing completeness and accuracy of data. |  |
| 11. **Analysis** | Page 6 and 7 |
| a. Qualitative and quantitative methods used to draw inferences from the data. |  |
| b. Methods for understanding variation within the data, including the effects of time as a variable. |  |
| 12. **Ethical considerations** - Ethical aspects of implementing and studying the intervention(s) and how they were addressed, including, but not limited to, formal ethics review and potential conflict(s) of interest. | Page 4 |
| **Results: What did you find?** |  |
| 13. **Results** | Page 7 and 8 and Figure 3 |
| a. Initial steps of the intervention(s) and their evolution over time (eg, time-line diagram, flow chart or table), including modifications made to the intervention during the project. |  |
| b. Details of the process measures and outcomes. |  |
| c. Contextual elements that interacted with the intervention(s). |  |
| d. Observed associations between outcomes, interventions and relevant contextual elements. |  |
| e. Unintended consequences such as unexpected benefits, problems, failures or costs associated with the intervention(s). |  |
| f. Details about missing data. |  |
| **Discussion: What does it mean?** |  |
| 14. **Summary** | Page 8 |
| a. Key findings, including relevance to the rationale and specific aims. |  |
| b. Particular strengths of the project. |  |
| 15. **Interpretation** | Page 9 and 10 |
| a. Nature of the association between the intervention(s) and the outcomes. |  |
| b. Comparison of results with findings from other publications. |  |
| c. Impact of the project on people and systems. |  |
| d. Reasons for any differences between observed and anticipated outcomes, including the influence of context. |  |
| e. Costs and strategic trade-offs, including opportunity costs. |  |
| 16. **Limitations** | Page 10 |
| a. Limits to the generalisability of the work. |  |
| b. Factors that might have limited internal validity such as confounding, bias or imprecision in the design, methods, measurement or analysis. |  |
| c. Efforts made to minimise and adjust for limitations. |  |
| **Conclusions** | Page 10 |
| a. Usefulness of the work. |  |
| b. Sustainability. |  |
| c. Potential for spread to other contexts. |  |
| d. Implications for practice and for further study in the field. |  |
| e. Suggested next steps. |  |
| **Other information** | Page 11 |
| 18. **Funding** - Sources of funding that supported this work. Role, if any, of the funding organisation in the design, implementation, interpretation and reporting. |  |
| *Ogrinc G, et al. BMJ Qual Saf 2015;0:1–7. doi:10.1136/bmjqs-2015-004411* |  |
| *Downloaded from http://qualitysafety.bmj.com/ on January 2, 2017* |  |
