## Appendix 1, 2 and 3. for "Evaluating a novel, integrative dashboard for health professionals’ performance in managing deteriorating patients: *quality improvement project*"

1- Table1. EWS documentation Status Summary for all observations taken in each hospital in Barts Health Trust.
2- Dashboard development stages
3- Interview questions.

### 1. Dashboard development stages

1- Developing began with initially creating a simple Vitals data table
2- A broader generation of vitals data is added.
3- Transforming the table into a thorough and more robust data visualisation of NEWS2, assessment and escalation of deteriorating patients via Qlik Sins.
4- Data of around 1.2 million recordings of 110,000 admissions from August to October 2020 were extracted from the Datawarehouse of Barts trust hospitals; pulled from EHR (Cerner Millennium®).
5- The dashboard metrics are indicators of the status of patients who needed escalation of care, such as vital signs and sepsis scoring and time of entry and by whom.
6- Post-development, data are pulled continuously until the present time. Dashboards were produced using SQL and final views were developed using a QVD table in the Qlik Sense® Server.
7- Validation of data was done by NK by evaluating 100 metrics accuracy and independently screened by NK and a quality officer.
8- Front end screening was conducted by nursing informatics to check the validity of presented data.
9- The dashboard was approved by the informatics lead, quality improvement and chief nursing information officer.

**2- Table 1.** EWS documentation Status Summary for all observations taken in each hospital in Barts Health Trust.

| Site Summary | | | | | | | | | | | | | | | |
|---|---|---|---|---|---|---|---|---|---|---|---|---|---|---|---|
| | Sep-19 | | | Oct-19 | | | Nov-19 | | | Dec-19 | | | Jan-20 | | |
| Location | Complete | Incomplete | % Complete Status | Complete | Incomplete | % Complete Status | Complete | Incomplete | % Complete Status | Complete | Incomplete | % Complete Status | Complete | Incomplete | % Complete Status |
| Newham University Hospital | 872 | 229 | 79% | 1147 | 319 | 78% | 9995 | 6493 | 61% | 32418 | 16017 | 67% | 19127 | 8387 | 70% |
| Royal London Hospital | 9679 | 1917 | 83% | 13498 | 3148 | 81% | 57585 | 19174 | 75% | 73852 | 21654 | 77% | 41128 | 11012 | 79% |
| St Bartholomew's Hospital | 6047 | 961 | 86% | 10213 | 1521 | 87% | 15747 | 2859 | 85% | 27804 | 5486 | 84% | 15668 | 2249 | 87% |
| Whipps Cross Hospital | 3308 | 241 | 93% | 33882 | 6835 | 83% | 62051 | 9231 | 87% | 66052 | 7860 | 89% | 37593 | 3858 | 91% |
| **Grand Total** | **19906** | **3348** | **86%** | **58740** | **11823** | **83%** | **145378** | **37757** | **79%** | **200126** | **51017** | **80%** | **113516** | **25506** | 82% |

#### 3- Interview questions

Introduction

Deteriorating dashboard is an EHR-integrated tool to audit the completion and percentage of staff recording of the assessment when patients deteriorating. Including vital signs, patients with high NEWS2, sepsis assessment by nurses and doctors, and sepsis 6 prescribing.

We would like to evaluate this tool as a QI project. This is the first cycle.
We are assessing the performance in numbers as showing in the dashboard as shown in historical view and interviewing some staff who utilise it to give us their opinion.
we hope after this assessment we will conduct any improvement needed, then re-evaluate in a few months.

Questions:

1. What is your job role?
2. How do you utilise the dashboard in your work?
3. Was there a training or orientation for you to use the dashboard?, how was it?
4. Is there an induction or training for current or future users?
5. What was your expectation of the dashboard? Were they met?
6. How beneficial is the dashboard for the purpose it's designed for? How valuable?
7. The dashboard is meant to audit decline or improvement in assessment, if detected, how well was this handled or coordinated?
8. Do you notice an improvement that took place? How or where?
9. If so, why do you think this change happened?
10. Did you notice any deterioration in the performance of staff?
11. If so, why do you think this happened?
12. Out of the data you got from the dashboard, is there any plan for improvement in staff practice or work flow strategy as a result of the dashboard?
13. -What are the downsides or negative aspects of the dashboard that needs to be improved?
14. -Is there anything that you would like to add to the dashboard?
15. If so, what would it be?
